# Plasma protein biomarkers predict both the development of persistent autoantibodies and type 1 diabetes 6 months prior to the onset of autoimmunity: the TEDDY Study

**DOI:** 10.1101/2022.12.07.22283187

**Authors:** Ernesto S. Nakayasu, Lisa M. Bramer, Charles Ansong, Athena A. Schepmoes, Thomas L. Fillmore, Marina A. Gritsenko, Therese R. Clauss, Yuqian Gao, Paul D. Piehowski, Bryan A. Stanfill, Dave W. Engel, Daniel J. Orton, Ronald J. Moore, Wei-Jun Qian, Salvatore Sechi, Brigitte I. Frohnert, Jorma Toppari, Anette-G. Ziegler, Åke Lernmark, William Hagopian, Beena Akolkar, Richard D. Smith, Marian J. Rewers, Bobbie-Jo M. Webb-Robertson, Thomas O. Metz, the TEDDY Study Group

## Abstract

Type 1 diabetes (T1D) results from an autoimmune destruction of pancreatic β cells. A significant gap in understanding the disease cause is the lack of predictive biomarkers for each of its developmental stages. Here, we conducted a blinded, two-phase case-control plasma proteomics analysis of children enrolled in the TEDDY study to identify biomarkers predictive of autoimmunity and T1D development. First, we performed untargeted proteomics analyses of 2,252 samples from 184 individuals and identified 376 regulated proteins. Complement/coagulation, inflammatory signaling and metabolic proteins were regulated even prior to autoimmunity onset. Extracellular matrix proteins and antigen presentation were differentially regulated in individuals with autoimmunity who progressed to T1D versus those who maintained normoglycemia. We then performed targeted proteomics measurements of 167 proteins in 6,426 samples from 990 individuals and validated 83 biomarkers. A machine learning analysis predicted both the development of persistent autoantibodies and T1D onset 6 months before autoimmunity initiation, with an area under the receiver operating characteristic curve of 0.871 and 0.918, respectively. Our study identified and validated biomarkers highlighting pathways affected in different stages of T1D development.

## Introduction

Type 1 diabetes (T1D) is a chronic metabolic condition that affects approximately 20 million people worldwide. Its associated morbidities (e.g., cardiovascular disease, blindness, and kidney failure) reduce life expectancy of individuals by 11 years^1^, and there is no cure yet for this disease. T1D results from a gradual destruction of insulin-producing β cells by an autoimmune response, which is associated with the appearance of autoantibodies against pancreatic islet proteins (hereafter referred to as ‘seroconversion’)^2,3^. However, the cause(s) that trigger and mechanisms that govern this autoimmune response are still poorly understood. The Environmental Determinants of Diabetes in the Young (TEDDY) study has an ambitious goal of identifying factors that contribute to β-cell autoimmunity or T1D, towards enabling the development of therapeutic interventions^4^. A key bottleneck in this process is the lack biomarkers that can accurately predict each step of T1D development.

Plasma proteomics analysis is a promising approach for discovering protein biomarkers^5-7^, and it has been applied to identify biomarkers of T1D onset^8-11^. Proteomics analysis can also provide important insights on the mechanism(s) of disease. Despite previous efforts^10,11^, there is still an urgent need for biomarkers that can predict the different stages of T1D development. Islet autoantibodies are excellent biomarkers and multi-positivity to islet autoantibodies predicts an almost inevitable development of T1D. However, there is a desperate need for biomarkers that predict and can be used to monitor the onset of islet autoimmunity. Moreover, it is also important to be able to distinguish between individuals that develop T1D vs. individuals that develop islet autoimmunity but not hyperglycemia, in order to appropriately focus potential treatments to the relevant stage of disease development.

Biomarker development is a long process, and many studies fall short due to the lack of systematic validation of candidates^12^. Here we conducted a robust T1D plasma protein biomarker discovery and validation study^13^ in the TEDDY cohort. We performed machine learning analysis to identify biomarker panels that can predict the development of both persistent autoantibodies and T1D with high accuracy and as early as 6 months before the appearance of the autoimmune response. By comparing with previously published proteomics models of insulitis using human islets and cultured β cells treated with cytokines, our results also provide insights on the mechanism of T1D development.

## Results

### Experimental design and discovery phase analysis

The study was based on a nested case-control design^4^ and aimed to identify biomarkers predictive of autoimmunity and T1D development, with samples divided into 8 groups: pre- and post-seroconversion for individuals that developed T1D or had persistent autoantibodies but with normoglycemia, each paired with respective control groups. The following comparisons were considered: I1: cases versus controls at pre-seroconversion with normoglycemia endpoint; T1: cases versus controls at pre-seroconversion with hyperglycemia endpoint; I2: cases versus controls at post-seroconversion with normoglycemia endpoint; T2: cases versus controls at post-seroconversion with hyperglycemia endpoint; I3: pre- vs post-seroconversion of cases with normoglycemia endpoint; and T3: pre- vs post-seroconversion of cases with hyperglycemia endpoint (**Figure 1**).

**Figure 1.**
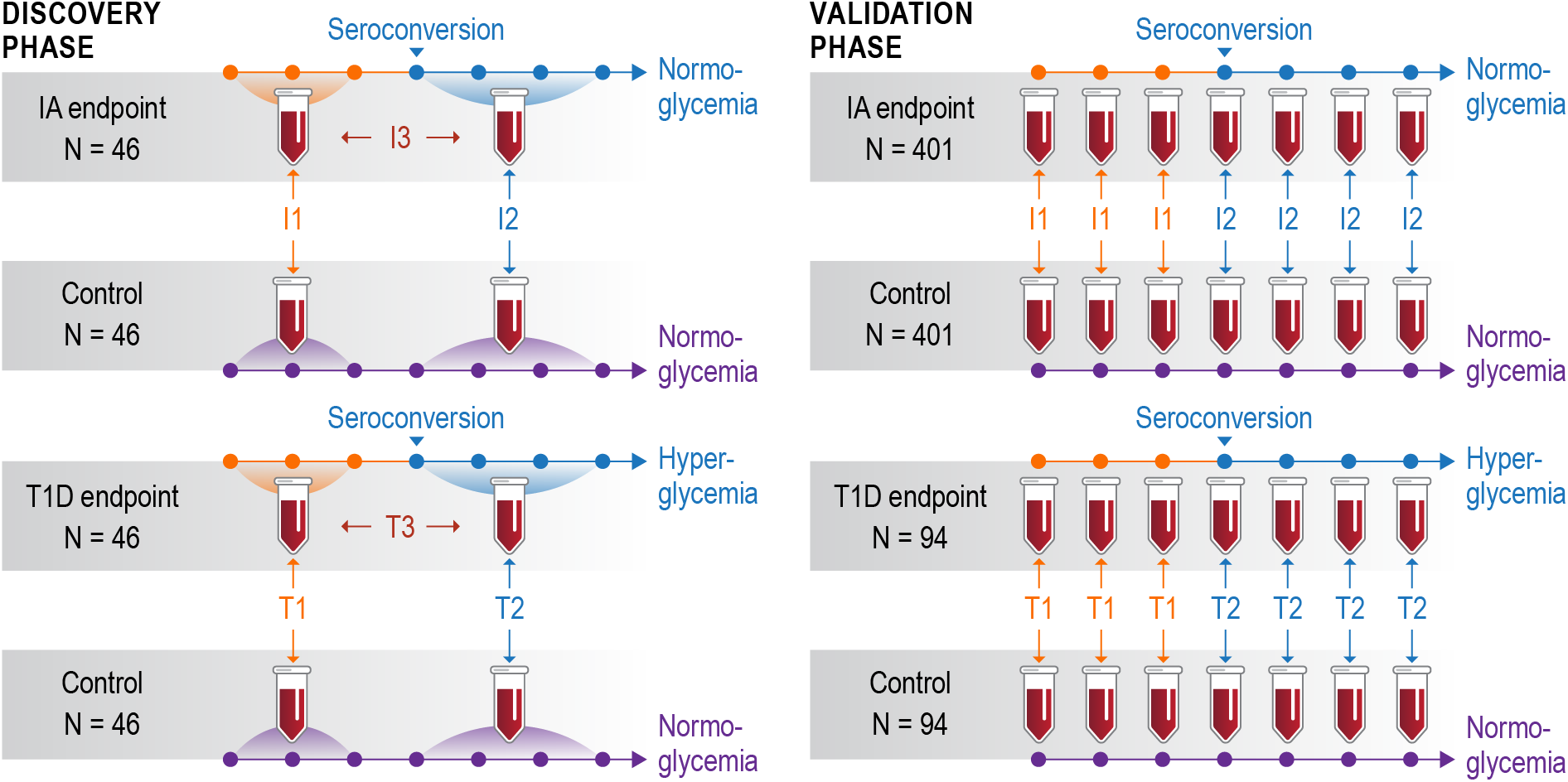
Study design: a two-phase study design to discover and validate biomarkers in human blood plasma. Individual plasma samples from a smaller number (N) of individuals were pooled from pre- and post-seroconversion visits and analyzed by in-depth untargeted proteomics in the discovery phase (left panel). Individual plasma samples from several collection time points (represented by the dots in the timeline) were analyzed in a larger cohort by targeted proteomics in the validation phase (right panel). Comparison I1: time point(s) before seroconversion in individuals that had normoglycemia (IA: islet autoimmunity with normoglycemia) at the end of the study paired against controls. Comparison T1: time point(s) before seroconversion in individuals that developed hyperglycemia (T1D: type 1 diabetes) paired against matched controls. Comparisons I2 and T2 have the same group of individuals as I1 and T1, respectively, but after seroconversion. Comparisons I3 and T3 compare individuals that remained normoglycemic or developed hyperglycemia before vs after seroconversion, respectively.

The study was comprised of two phases: a *discovery phase* focused on a deep proteomics analysis of pooled samples from a limited number of individuals^5^ (N = 184); and a subsequent *validation phase* with selected biomarker candidates analyzed by targeted proteomics in many samples from a much larger cohort^14^ (N = 990) across multiple time points (**Figure 1**). The characteristics and demographic information for both discovery and validation phase cohorts are presented in **Table 1**. A total of 1488 mass spectrometry analyses from 62 multiplexed proteomics sets were performed in the discovery phase. To ensure quality across 18 months of data collection, we developed and implemented an automated quality control system named QC-ART (Quality Control Analysis in Real Time)^15^. This tight quality control analysis assured that consistent data were collected across the study. The data profile had very similar distributions of peptide abundances across different multiplexed sets (**Figure S1A**) and number of identified peptides in each group (**Figure S1B**). A total of 36,252 peptides derived from 1,720 proteins were identified and after normalizing to a reference sample that was included in each multiplexed proteomics set, peptides were sequentially removed from the data set based on the following criteria: (I) detected in 2 or fewer samples across any group, (II) coefficient of variance greater than 150%, (III) detected in fewer than 2 matched case-control pairs, and (IV) *p*-value > 0.05 across different comparisons. These criteria resulted in a final discovery phase proteomics dataset that included 376 significant proteins (373 with ≥ 2 peptides and 3 with 1 peptide) at a *p*-value threshold of ≤0.05 (**Figure 2A, Table S1**).

**Table 1.** Characteristics of the study cohort. Abbreviations: FDR, first-degree relative; GADA, glutamic acid decarboxylase autoantibody, GP, general population; HLA, human leukocyte antigen; IA, islet autoimmunity with normoglycemia; IA-2A, islet antigen-2 autoantibody; IAA, insulin autoantibody; T1D, type 1 diabetes.

|  |  | Discovery |  |  |  | Validation |  |  |  |
| --- | --- | --- | --- | --- | --- | --- | --- | --- | --- |
|  |  | T1D |  | IA |  | T1D |  | IA |  |
|  |  | Cases | Matched Controls | Cases | Matched Controls | Cases | Matched Controls | Cases | Matched Controls |
| **Number** |  | 46 | 46 | 46 | 46 | 94 | 94 | 401 | 401 |
| **Case Seroconversion Age (Months)** | **Median** | 12 | . | 23 | . | 12 | . | 22 | . |
|  | **Q1** | 9 | . | 14 | . | 10 | . | 12 | . |
|  | **Q3** | 18 | . | 33 | . | 19 | . | 33 | . |
| **Sex** | **Female** | 25 | 25 | 17 | 17 | 43 | 43 | 179 | 179 |
|  | **Male** | 21 | 21 | 29 | 29 | 51 | 51 | 222 | 222 |
| **Clinical Center** | **Colorado** | 8 | 8 | 4 | 4 | 13 | 13 | 57 | 57 |
|  | **Georgia/Florida** | . | . | 1 | 1 | 6 | 6 | 28 | 28 |
|  | **Washington** | 4 | 4 | 5 | 5 | 6 | 6 | 37 | 37 |
|  | **Finland** | 23 | 23 | 23 | 23 | 31 | 31 | 113 | 113 |
|  | **Germany** | 6 | 6 | 1 | 1 | 13 | 13 | 33 | 33 |
|  | **Sweden** | 5 | 5 | 12 | 12 | 25 | 25 | 133 | 133 |
| **HLA-DR-DQ Genotypes** | **HLA Ineligible** | 1 | 1 | . | . | 1 | 3 | 1 | 2 |
|  | **DR3/4** | 26 | 20 | 25 | 17 | 55 | 39 | 211 | 152 |
|  | **DR4/4** | 7 | 6 | 10 | 9 | 13 | 18 | 65 | 71 |
|  | **DR4/8** | 7 | 5 | 8 | 6 | 14 | 10 | 61 | 67 |
|  | **DR3/3** | 1 | 6 | 3 | 11 | 5 | 9 | 46 | 81 |
|  | **FDR-specific** | 4 | 8 | . | 3 | 6 | 13 | 17 | 28 |
|  |  | T1D |  | IA |  | T1D |  | IA |  |
|  |  | Cases | Matched Controls | Cases | Matched Controls | Cases | Matched Controls | Cases | Matched Controls |
| Family History of T1D | GP | 28 | 28 | 42 | 42 | 61 | 61 | 309 | 309 |
|  | FDR: mother | 3 | 11 | 1 | 3 | 4 | 15 | 16 | 37 |
|  | FDR: father | 11 | 5 | 2 | 1 | 20 | 14 | 54 | 40 |
|  | FDR: both parents | . | . | . | . | 1 | . | 1 | . |
|  | FDR: sibling | 4 | 2 | 1 | . | 8 | 4 | 21 | 15 |
| Type of First Autoantibody | Not IA+ | . | 44 | . | 45 | . | 89 | . | 392 |
|  | IAA only | 27 | 2 | 27 | . | 53 | 4 | 194 | 0 |
|  | GADA only | 8 | . | 11 | 1 | 14 | 1 | 132 | 3 |
|  | IA-2A only | . | . | 1 | . | . | . | 6 | . |
|  | Two or more autoantibodies | 11 | . | 7 | . | 27 | . | 69 | . |

**Figure 2.**
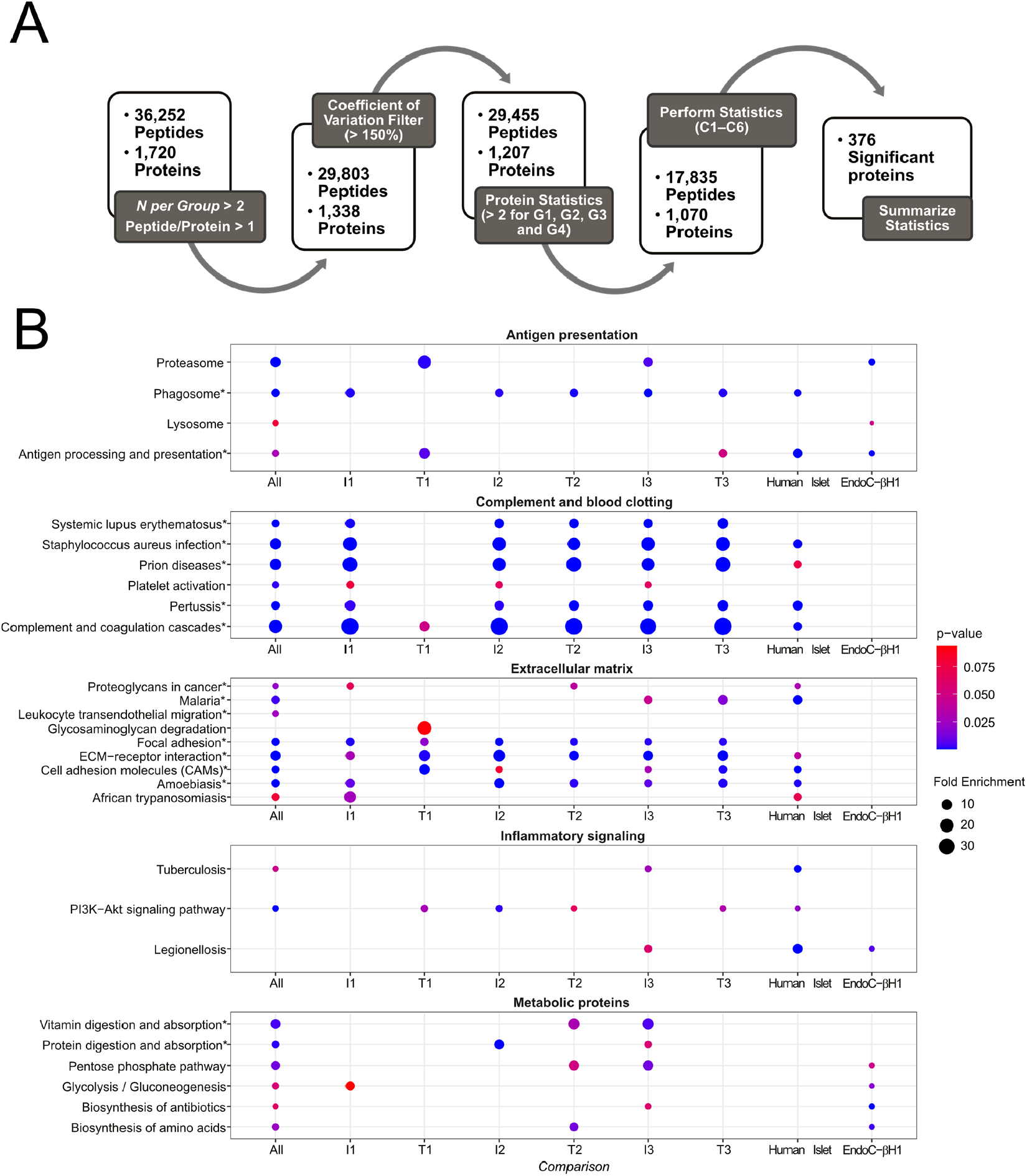
Discovery phase data analysis. (A) Discovery phase data quality and statistical analysis workflow. Sequential pre-filtering steps focused on identification and removal of high variability (steps 1-2) and low coverage (step 3) peptides and proteins. Resulting proteins and peptides were submitted to multiple statistical comparisons in the context of autoimmunity and T1D development (steps 4-5). (B) Functional-enrichment analysis. The 376 differentially abundant proteins identified from the discovery phase were submitted to function-enrichment analysis with DAVID, using the KEGG annotation. Pathways were plotted as circles with sizes based on their fold enrichment and colors based on p-values. Individual pathways were grouped into larger biological processes based on the overlapping proteins between each pathway. Pathways that were also enriched among the 167 targets of the validation phase are marked with asterisks. Comparison I1: time point(s) before seroconversion in individuals that had normoglycemia at the end of the study paired against controls. Comparison T1: time point(s) before seroconversion in individuals that developed hyperglycemia paired against matched controls. Comparisons I2 and T2 have the same group of individuals as I1 and T1, respectively, but after seroconversion. Comparisons I3 and T3 compare individuals that remained normoglycemic or developed hyperglycemia before vs after seroconversion, respectively.

### Biological pathways regulated in islet autoimmunity and T1D development

A functional-enrichment analysis of the discovery phase data showed that 22 pathways were overrepresented among the 376 differentially abundant proteins and their proteoforms (**Figure 2B**). To facilitate the interpretation, we further grouped these pathways into fewer biological processes based on the components of each pathway that were regulated in the different comparisons. We plot the pathways as circles with their size being proportional to the fold enrichment and colored based on the enrichment significance (**Figure 2B**). Complement and blood clotting, antigen presentation, extracellular matrix, nutrient digestion and absorption, cellular metabolism and inflammatory signaling processes were significantly enriched with differentially abundant proteins (**Figure 2B**). We compared the functional-enrichment analysis of the TEDDY proteomics data to published proteomics analyses of human islets^16^ and the β-cell line EndoC-βH1^17^ from the Human Islet Research Network (HIRN). Each sample type was treated with pro-inflammatory cytokines IL-1β+IFNγ as a model of insulitis. Proteins related to complement and blood clotting, antigen presentation, extracellular matrix, and inflammatory signaling were also enriched among the IL-1β+IFNγ regulated proteins in the human islet study (**Figure 2B**). In EndoC-βH1 cells, pathways related to antigen presentation, inflammatory signaling and cell metabolism were regulated similarly to the plasma signatures (**Figure 2B**). This shows that similar inflammatory signatures that occur in islets treated with pro-inflammatory cytokines can also be detected in plasma of individuals during T1D development.

#### Extracellular matrix

Pathways related to the extracellular matrix were commonly enriched among the different comparisons. However, there was only a small overlap of significant proteins between the different comparisons as shown in the heatmap. At pre-seroconversion, individuals who only developed autoimmunity but remained normoglycemic had 14 regulated proteins (12 upregulated) (Comparison I1, **Figure 3**), while individuals that developed hyperglycemia had 10 regulated proteins (all upregulated) (Comparison T1, **Figure 3**). Post-seroconversion, the scenario became more distinct, with the islet autoimmunity with normoglycemia group having 24 out of 25 regulated proteins downregulated, while the group that developed hyperglycemia had 17 out of the 17 regulated proteins upregulated (Comparisons I2 and T2, respectively, **Figure 3**).

**Figure 3.**
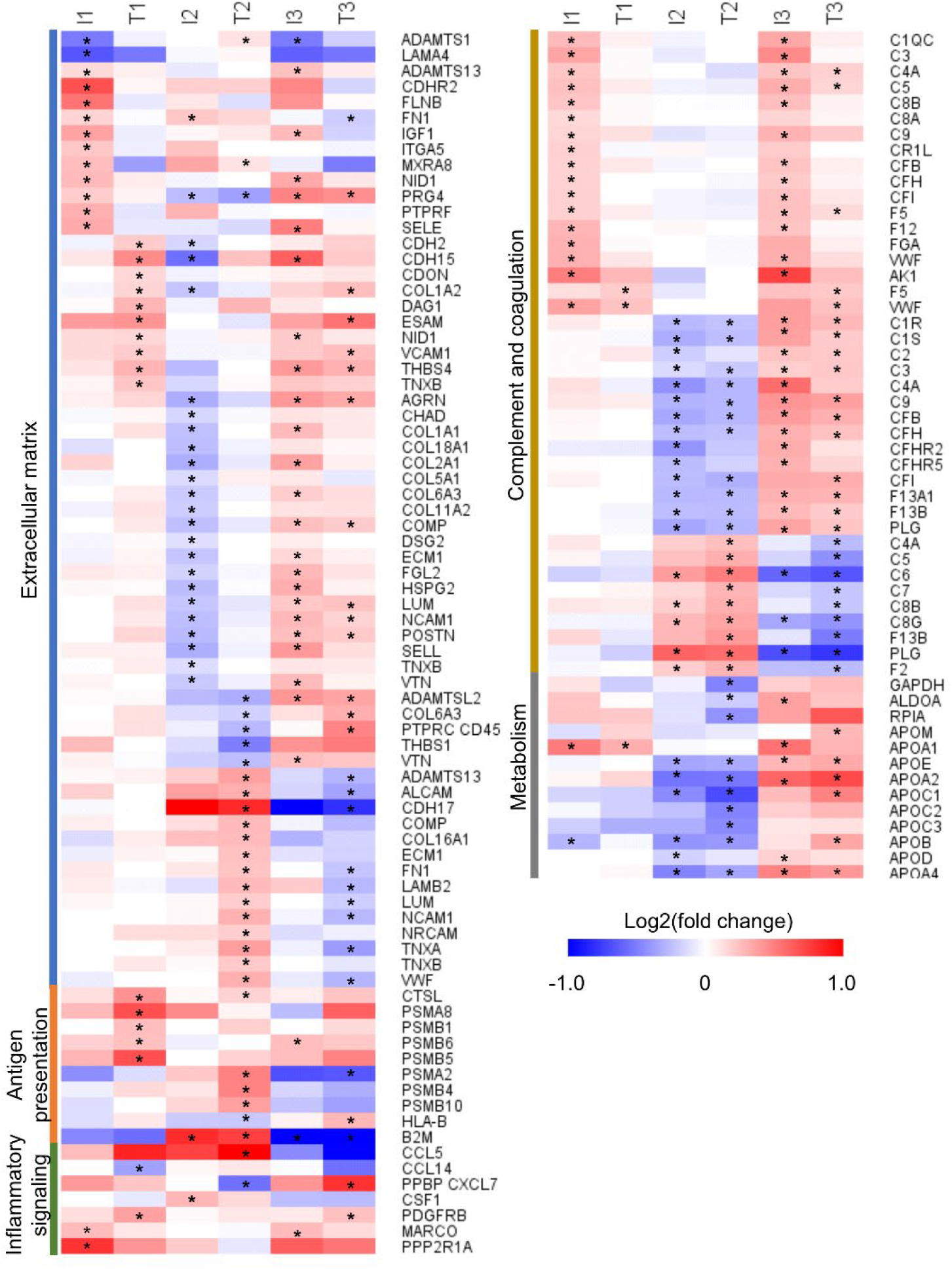
Regulated pathways. The 376 differentially abundant proteins identified from the discovery phase were submitted to function-enrichment analysis with DAVID, using the KEGG annotation. The heatmap shows the proteins enriched for each pathway. Asterisks mark those proteins in specific comparisons that were statistically significant. Comparison I1: time point(s) before seroconversion in individuals that had normoglycemia at the end of the study paired against controls. Comparison T1: time point(s) before seroconversion in individuals that developed hyperglycemia paired against matched controls. Comparisons I2 and T2 have the same group of individuals as I1 and T1, respectively, but after seroconversion. Comparisons I3 and T3 compare individuals that remained normoglycemic or developed hyperglycemia before vs after seroconversion, respectively. Proteins are named based on Uniprot gene names.

#### Antigen presentation

Antigen processing and presentation was the most distinctive pathway at the pre-seroconversion time point when comparing the group that developed T1D vs those who only displayed autoimmunity with normoglycemia. In the group that developed T1D, a higher level of antigen-processing proteins was observed, including cathepsin L1 (CTSL) (protein names are abbreviated using their Uniprot gene names) and proteasome subunits PSMA8, PSMB1, PSBM5 and PSBM6 (Comparison T1, **Figure 3**). Cathepsin L1 and proteasome subunits PSMA2, PSBM4 and PSBM10 were also higher after seroconversion, but were accompanied also by the antigen presenting complex HLA class I (HLA-B) and β-2-macroblobulin (B2M) (Comparison T2, **Figure 3**).

#### Inflammatory signaling

Four cytokines and chemokines were regulated across different comparisons. C-C motif chemokine 14 (CCL14) was downregulated pre-seroconversion in the T1D group compared to the control (Comparison T1, **Figure 3**). C-C motif chemokine 5 (CCL5) and proplatelet basic protein (PPBP or CXCL7) were up- and down-regulated, respectively, in the T1D group compared to the control at the post-seroconversion time point (Comparison T2, **Figure 3**). Receptors, such as platelet-derived growth factor receptor beta (PDGFRB) and macrophage receptor MARCO, and signaling transduction proteins, such as serine/threonine-protein phosphatase 2A 65 kDa regulatory subunit A alpha isoform (PPP2R1A), were also regulated (**Figure 3**).

#### Complement and coagulation

Complement factors C1QC, C3 C4A, C5, C8A, C8B, C9, CR1L, CFB, CFH and CFI - and coagulation factors F5, F12, fibrinogen α and γ, von Willebrand and adenylate kinase - were higher at the pre-seroconversion time point in plasma of individuals with islet autoimmunity with normoglycemia endpoint vs. respective controls (Comparison I1, **Figure 3**). Proteoforms of F5 and von Willebrand factors were upregulated in the individuals that developed T1D at the same early time point (Comparison T1, **Figure 3**). Post-seroconversion, both groups had lower levels of most coagulation and complement factors compared to their respective controls. However, specific proteoforms were regulated in the opposite way (Comparisons I2 and T2, **Figure 3**), probably reflecting processing or post-translational modifications of these proteins.

#### Metabolic proteins

Among the central carbon metabolism enzymes, glyceraldehyde-3-phosphate dehydrogenase (GAPDH), fructose-bisphosphate aldolase A, and ribose-5-phosphate isomerase were reduced post-seroconversion in the group that developed T1D but not in the islet autoimmunity with normoglycemia group (Comparisons I2 and T2, **Figure 3**), suggesting an abnormal sugar metabolism. Lipoproteins represent another class of metabolic proteins regulated in plasma. Apolipoprotein (Apo) A1 was increased in both cohorts of individuals that developed T1D and islet autoimmunity with normoglycemia pre-seroconversion but had similar levels to the control after seroconversion (**Figure 3**). Conversely, Apo A2, A4, B, C1, C2, C3, D, E, H and J had similar levels compared to the controls in both groups pre-seroconversion but declined after seroconversion (**Figure 3**). Overall, these data indicate changes in metabolic proteins that precede hyperglycemia.

### Validation of protein biomarker candidates

We performed a systematic prioritization of the candidate biomarkers from the discovery phase based on the following criteria: (I) statistical significance at Benjamini-Hochberg adjusted *p*-value ≤0.05, (II) ≥ 2 peptides identified per protein, a spectral count (SpC) ≥ 20 and unadjusted p-value < 0.005, (III) ≥ 2 peptides identified per protein, SpC ≥ 20, detected in more than 23 samples, and machine learning (ML) to determine the group of proteins that are the most predictive of each of the 6 comparisons, or (IV) unadjusted p-value ≤0.05 but were previously described as potential T1D onset biomarkers in the literature^8-11^ (**Figure 4A**). This analysis led to the selection of 167 proteins for the validation phase, of which 811 peptides were selected for targeted proteomics assay development (as described in methods). Similar to the discovery phase, we developed an informatics tool named Q4SRM (Quality control analysis for Selected Reaction Monitoring)^18^ to systematically track data quality across 29 months of analyses. A total of 694 peptides from all 167 proteins were successfully monitored until the end of the study (**Figure 4B and Table S2**). An additional post-hoc quality control analysis showed a strikingly high correlation (>95%) for almost all the 6,426 targeted proteomics analyses performed (**Figure S2**). From the measured peptides, 127 peptides from 83 (50%) proteins were significant and showed similar abundance patterns to the discovery phase across comparisons I1, T1, I2, and T2 validating them as biomarkers (**Figure 4** and **Table S3-S4**). The 83 validated proteins belong to all major biological processes observed as regulated in T1D development in the discovery phase: antigen presentation, complement and blood clotting, extracellular matrix, inflammatory signaling and metabolic proteins.

**Figure 4.**
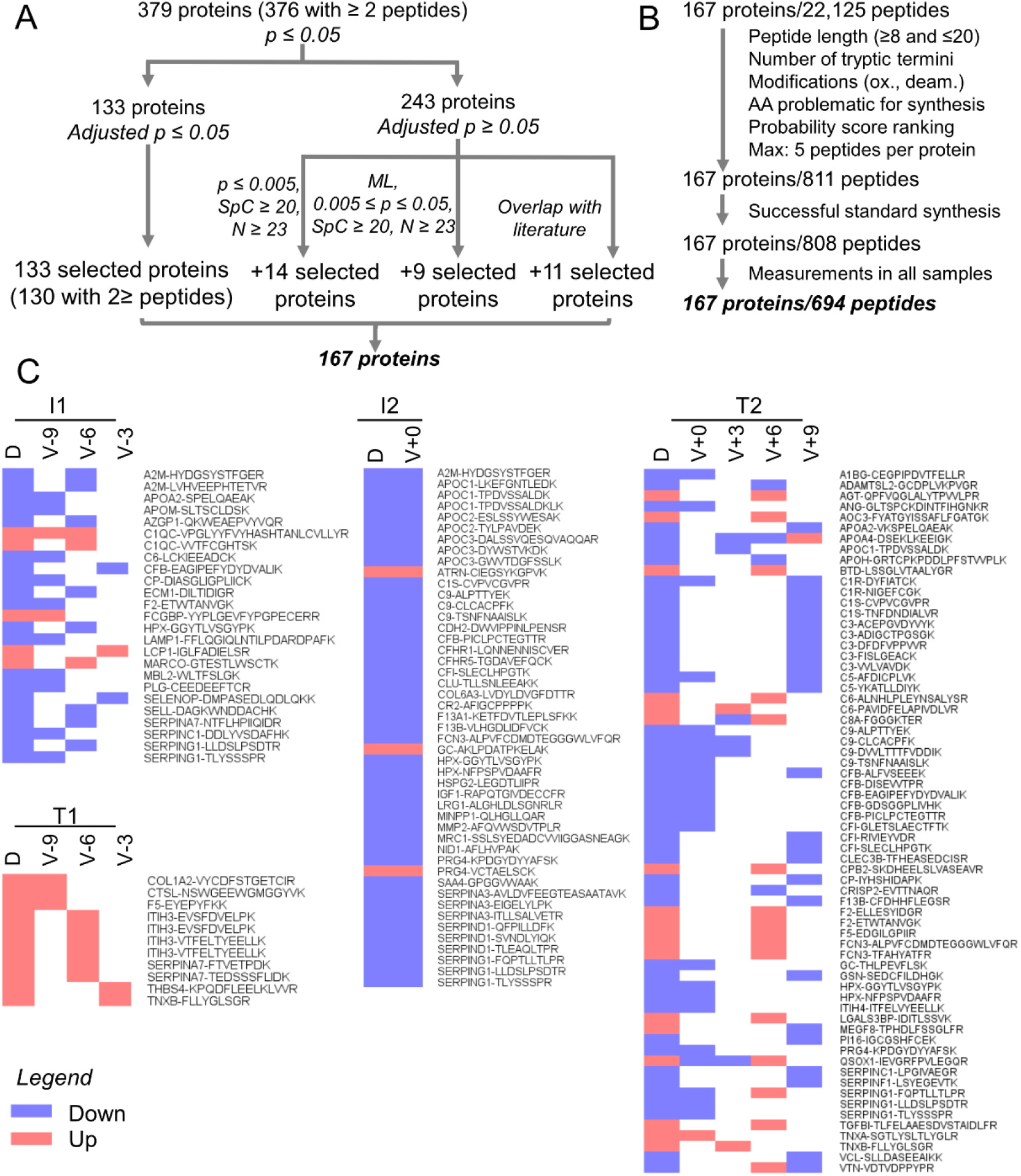
Validation phase data analysis. (A) Biomarker candidates were selected first based on statistical test with *p*-value correction. Additional candidates were selected for validation based on the p-value, number of samples in which the peptide was detected, spectral count, machine learning and previous reports in the literature. (B) Up to 5 peptides for each candidate protein were selected based on their physicochemical properties and probability ranking for a likely successful measurement by targeted proteomics. (C) Cross-validated proteins across discovery (D) and validation (V) phases. Only significant, validated proteins are represented in the heatmap and colored based on their regulation. Time points are represented by months prior (-) or post (+) seroconversion. Comparison I1: time point(s) before seroconversion in individuals that had normoglycemia at the end of the study paired against controls. Comparison T1: time point(s) before seroconversion in individuals that developed hyperglycemia paired against matched controls. Comparisons I2 and T2 have the same group of individuals as I1 and T1, respectively, but after seroconversion. Proteins are named based on Uniprot gene names followed by the peptide sequence.

### Machine learning models for predicting T1D onset

Machine learning is a powerful approach to identify individual or combinations of biomarkers that can predict a phenotype. Therefore, we performed machine learning analysis to identify biomarkers that can predict the development of islet autoimmunity with normoglycemia or T1D prior to patient seroconversion. We used logistic regression with a LASSO penalization to build machine learning models that can predict the different outcomes. This analysis can identify models based on panels of peptides that best predict the different outcomes, and they were tested by cross-validation repeated for 100 bootstrap iterations. The receiver operating characteristic curves from this analysis show that both islet autoimmunity with normoglycemia and T1D onset can be predicted with high accuracy at 6 months prior to the seroconversion time with an average area under the curve of 0.871 and 0.918, respectively (**Figure 5A**). **Figure 5B** shows the proteins that correlate to the panel of peptides that were selected by the machine learning analysis to build the models. Among the most important proteins, i.e. the ones that appeared with more frequency across the training models, there were proteins from the complement and coagulation cascades (e.g. C4B, C5, C6, C8B, C9, F2 and F5), extracellular matrix (e.g. MMP2, COL1A1, COL1A2, WVF and ADAMTS13), and antigen processing and presentation (HLA-A, HLA-B and B2M) (**Figure 5B**), suggesting that they are important processes in the disease development. A total of 28 out of the 116 selected peptides were commonly selected across both I1 and T1 comparisons, while 81 were selected only in the I1 comparison and 7 only in T1 (**Figure 5B**), showing that both islet autoimmunity with normoglycemia or T1D development have some overlapping but also distinct signatures. Overall, the machine learning analysis showed that islet autoimmunity with normoglycemia or T1D development can be predicted even 6 months prior to the onset of islet autoimmunity.

**Figure 5.**
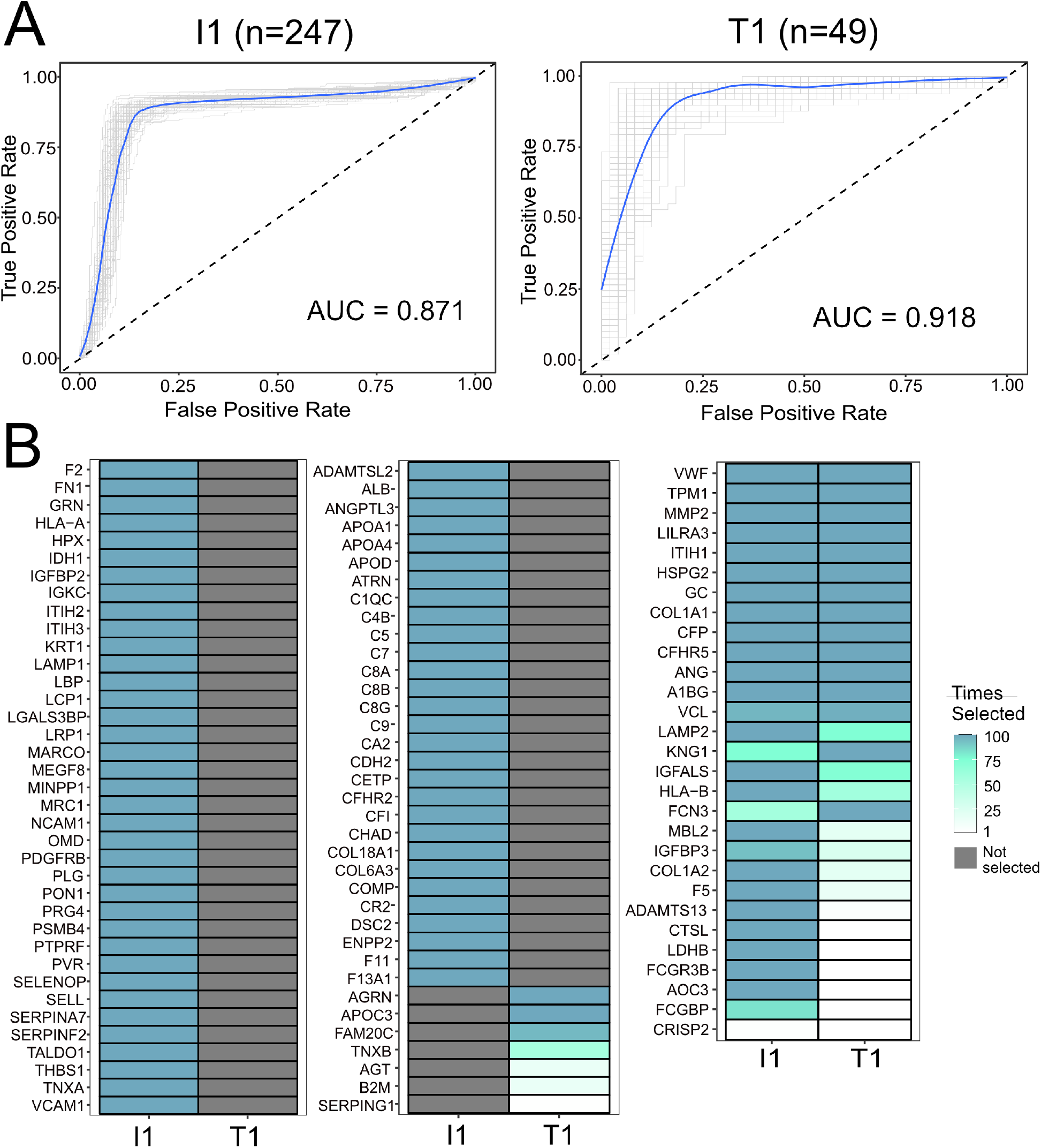
Prediction of autoimmunity with normoglycemia or type 1 diabetes (T1D) onset prior to seroconversion by machine learning analysis. (A) The panels show receiver operating characteristic (ROC) curves of peptide panels that predict normoglycemia (comparison I1) and T1D onset (comparison T1) at 6 months prior to the seroconversion. The numbers (n) of case-control pairs used at each time point are shown at the top of each ROC curve. Individual bootstrap curves are shown in gray with the mean curve given in blue. (B) Heatmaps showing the selected proteins and their frequencies of being kept in the model over the 100 bootstrap iterations for the most important peptide features used to predict the model. The left two panels contain proteins that were selected in only one comparison, whereas the right panel shows proteins that were commonly selected. Proteins are named based on Uniprot gene names.

## Discussion

We initially identified 376 differentially abundant proteins among the varying points of islet autoimmunity with normoglycemia and T1D development in a cohort of the TEDDY study. These proteins were overrepresented in processes related to T1D development such as complement and blood clotting, antigen presentation, extracellular matrix, nutrient digestion and absorption, cellular metabolism and inflammatory signaling (**Figure 6**). Importantly, these processes were also regulated in human islets and cultured β cells stimulated with pro-inflammatory cytokines to mimic the insulitis process. This suggests that some of these processes also occur in the pancreas during T1D development. Overall, our data showed a regulation in the complement and coagulation cascades. Polymorphism in complement has been associated with a higher risk of T1D development^19,20^. Increased complement activation and deposition have been shown in pancreata from individuals with T1D^21^. Patients with T1D also have increased clotting condition, including upregulation in platelet aggregation and coagulation activity and reduction in fibrinolysis^22^. Complement can also participate in opsonization of pathogens or dead cells towards phagocytosis (**Figure 6**).

**Figure 6.**
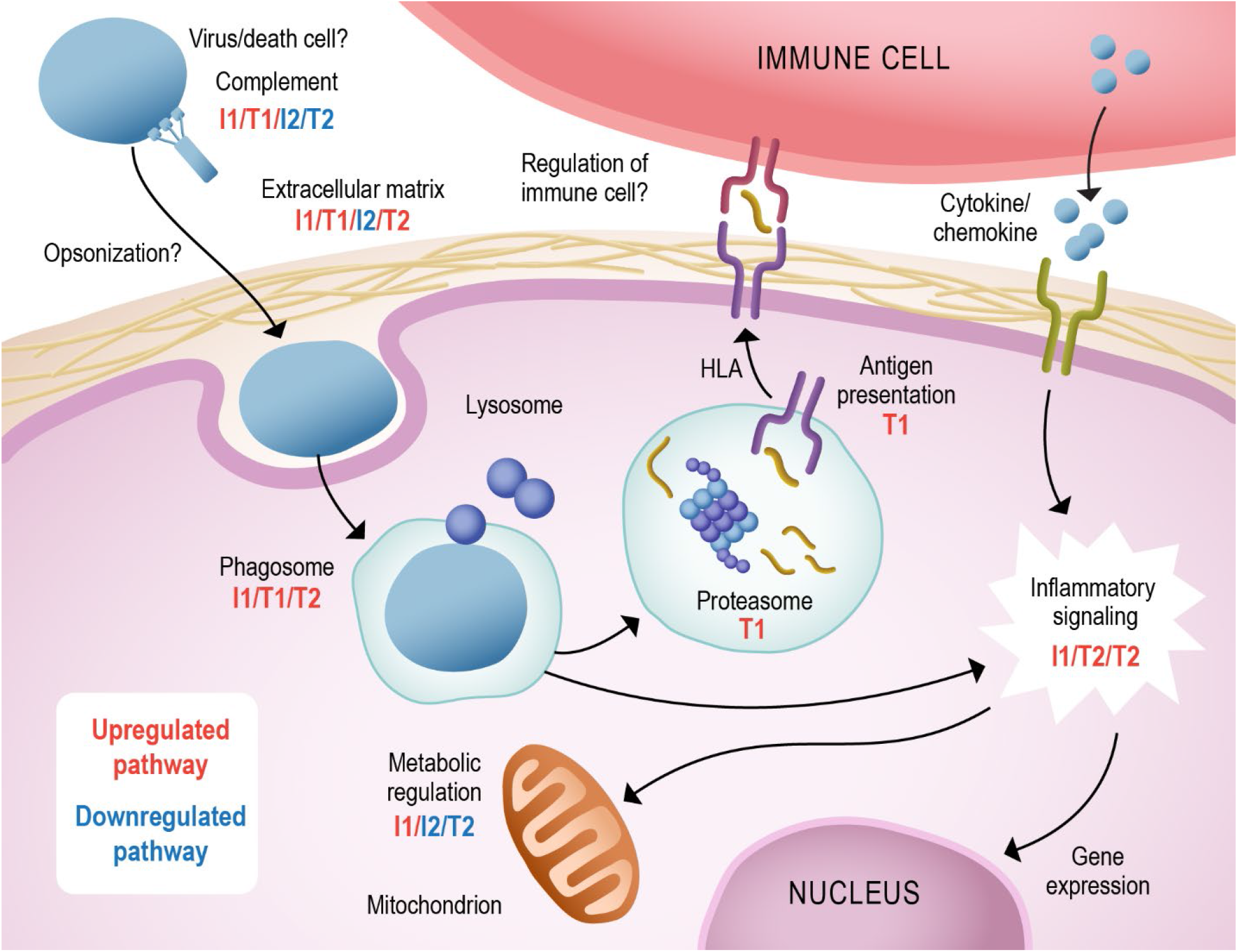
Summary of pathways regulated in autoimmunity and type 1 diabetes development. Many components of the complement cascade were found to be increased pre-seroconversion (comparisons T1/I1) and decreased post-seroconversion (T2/I2). An increase in phago/lysosome components was observed in comparisons T1/I1/T2. However, an increase in proteasome and antigen presentation components was only observed in T1. This process can trigger cellular signaling along with the stimulation of cytokine/chemokine receptors, regulating gene expression and cell metabolism (I1/T2/I2). We also observed a regulation of the extracellular matrix proteins (up in T1/I1/T2 and down in I2), which can regulate the interaction with immune cells. Comparison T1: time point before seroconversion in individuals that developed hyperglycemia paired against matched controls. Comparison I1: time point before seroconversion in individuals that had normoglycemia at the end of the study paired against controls. Comparisons T2 and I2 have the same group of individuals as T1 and I1, respectively, but after seroconversion.

The phagocytosis and lysosome components were also shown to be regulated in our data (**Figure 6**). This process is involved in pathogen and dead cell destruction and antigen presentation. The subsequent processes of proteasome antigen processing and presentation with HLA (human leukocyte antigen) were only upregulated at the pre-seroconversion stage of individuals that developed T1D (comparison T1) (**Figure 6**), reinforcing the importance of this process in the disease development. It is possible that higher proteasome levels result in abnormal antigen presentation and autoimmunity development. Polymorphism on antigen presentation gene HLA is indeed the major risk factor for developing T1D^23^. The HLA variants can differentially present islet self-antigens and are believed to be involved in autoimmunity development^24^. During islet autoimmunity, pro-inflammatory cytokines and chemokines are produced, triggering β-cell apoptosis and helping to recruit leukocytes and leading to insulitis^25^. This signaling also leads to regulation in gene expression and cell metabolism, which is observed in our data (**Figure 6**).

Our data show shifts in metabolic proteins even pre-seroconversion (**Figure 6**). Changes in metabolite profiles have been shown to predict development of autoantibodies 6 months prior to seroconversion^26^. In addition, metabolite profiles detected in 3–9-month-old children from the TEDDY study are predictive of their developing T1D by the age of 6 years^27^. In addition, abnormal proinsulin-to-C-peptide ratio can be detected 12 months prior to the onset of T1D^28^, suggesting a dysfunction in insulin processing that may affect the body metabolism even before causing hyperglycemia. Furthermore, several components of plasma lipoproteins were downregulated after seroconversion. Triacylglycerols, which are major components of plasma lipoproteins, have been shown to be lower in children that developed T1D compared to children that had islet autoimmunity but who remained normoglycemic^29^. Lipoprotein subunits, such as apolipoprotein CIII, have been linked to T1D development. Apolipoprotein CIII has been shown to trigger β-cell apoptosis^30^. Overall, changes in metabolism precede the disease onset and may also be involved in T1D development.

Another process highly regulated in our data was the extracellular matrix (**Figure 6**). Circulating extracellular matrix proteins are good indicators of tissue damage^31^, and may indicate damage on the pancreatic islets. In addition, during recruitment of leukocytes, the islet extracellular matrix undergoes major remodeling to allow cell infiltration^32^. Our data show a different profile on plasma extracellular proteins between the individuals with IA that developed T1D or had normoglycemia, possibly enabling or impeding β-cell destruction^33^.

In clinical diagnosis, T1D is diagnosed by blood glucose levels or by glycated hemoglobin^3^. For predictive biomarkers, HLA genotype and autoantibodies against islet proteins have been used but they lack enough discriminative power due to the heterogeneity of the disease^34^. Biomarkers based on T cells are currently being developed but require further validation^35^. Proteomics has been applied to identify T1D biomarkers, but some of these were focused on disease diagnosis after onset^8,9^. In biomarker studies prior to T1D onset, von Toerne et al. performed a proteomics discovery and validation study on samples from individuals after seroconversion to identify biomarkers that can diagnose the onset of islet autoimmunity and T1D development. They identified several circulating biomarkers of islet autoimmunity, and found that a protein panel composed by hepatocyte growth factor activator, complement factor H, ceruloplasmin and age can predict progression time to T1D^10^. Moulder et al. performed untargeted proteomics analysis in a longitudinal study from 3 months to 12 years of age for 13 individuals that developed T1D vs. age matched controls and found that the profile of proteins such as complement proteins and apolipoproteins can predict the onset of T1D^11^. Here, we performed a study to identify and validate biomarkers of different stages of the disease and the likelihood of developing T1D. Unlike the study by Moulder et al that matched case-control pairs based on age, we make our comparisons in relation to seroconversion. We identified and validated 83 biomarkers of islet autoimmunity and T1D development prior to the onset of the disease. Furthermore, we performed machine learning analysis and identified panels of proteins that can predict both the development of persistent autoantibodies with normoglycemia and T1D even 6 months prior to the appearance of the autoimmune response. We believe evaluation of these promising predictive protein panels in other ongoing prospective studies of development of autoimmunity and T1D in human cohorts could aide in the development of new prognostics and therapeutics.

One limitation of our study is that the validation was not performed in an independent cohort of samples. Validation in independent cohorts of samples can eliminate some confounding factors based on geographical and populational biases. However, our cohort includes individuals from 6 different centers in the US and Europe, which can reduce some of the regional confounding factors. Another limitation of our study is that the machine learning models were also not validated in an independent cohort of samples. However, they have gone through 100 bootstrap iterations of repeated cross-validation for the robustness of the analysis. Therefore, these two limitations are among the points that need to be further evaluated in additional studies of independent cohorts before implementing our findings in clinical practice. Despite these limitations, our results provided biological insights on the molecular pathways regulated in T1D development and identified biomarker candidates for the disease.

## Methods

### Study design, sample cohort, batching and randomization

The study was conducted after approval from the Institutional Review Boards of the University of South Florida (USF) and the Pacific Northwest National Laboratory (PNNL) in accordance with federal regulations. The study was designed to primarily identify biomarkers of pre-autoimmune response in children who progressed or not to T1D, and the following comparisons were considered: I1) cases versus controls at pre-seroconversion with normoglycemia endpoint, T1) cases versus controls at pre-seroconversion with hyperglycemia endpoint, I2) cases versus controls at post-seroconversion with normoglycemia endpoint, T2) cases versus controls at post-seroconversion with hyperglycemia endpoint, I3) pre- vs post-seroconversion of cases with normoglycemia endpoint, and T3) pre- vs post-seroconversion of cases with hyperglycemia endpoint (**Figure 1**).

TEDDY study participants have higher genetic risk of developing T1D, and to reduce the study to a manageable size, samples were previously matched based on clinical center, gender, and family history of T1D^4^. This nested case-control design resulted in 401 one-to-one pairs for the islet autoimmunity with normoglycemia endpoint and 94 pairs for the T1D endpoint^4^. The characteristics of the subset of case-control samples are listed in **Table 1**. The study was designed with a discovery and a validation phase to ensure an in-depth and robust analysis and was also conducted in a blinded fashion until the conclusion of the validation phase. Sample selection, batching, and randomization were performed at USF, whereas proteomics measurements were conducted at PNNL. Randomization was performed to assure that the study endpoints and patient time points were appropriately dispersed across the study and that the nested case-control pairs were analyzed within the same batch during processing to match the statistical design.

### Discovery Phase - Untargeted proteomics analysis

A statistical power analysis was performed to determine the number of case-control pairs needed in each study group, using a previous proteomics dataset consisting of 16,928 peptides measured in 12 individuals across multiple time points. Using the power.t.test function in R package (v3.2.3), it was determined that 23 case-control pairs were required to reach 80% power to detect a 2-fold difference utilizing a variance estimate associated with the 75-th percentile of measurements from the proteomics data. This number was doubled to 46 case-control pairs to account for untargeted proteomics missing data and resulted in 2252 plasma samples considering the multiple time points which were combined per donor within pre- or post-seroconversion into 368 samples due to costs and logistics. In the analysis, fourteen of the most abundant proteins in each sample were depleted using a Hu-14 4.6 × 100 mm MARS column (Agilent Technologies, Palo Alto, CA) coupled to a 1200 series HPLC (Agilent) and concentrated in Amicon centrifugal filters (3-kDa MWCO, Millipore, Burlington, MA). Proteins were digested in 96-well plates^36^, and peptides were labeled with 8-plex iTRAQ reagent (Applied Biosystems, Foster City, CA) following manufacturer recommendations. A pooled reference sample was created by mixing aliquots of each sample and was used for normalization across different datasets. The multiplexed iTRAQ-labeled samples were fractionated by high pH reversed phase chromatography and analyzed on a nanoAquity UPLC® system (Waters) connected to a LTQ Orbitrap Velos mass spectrometer (Thermo Scientific)^15,37^. Mass spectra were processed using Decon2LS_V2 and DTA Refinery^38,39^, with peptides identified using MSGF+^40^ by searching against the human SwissProt sequences of the Uniprot Knowledgebase. The parameters included: (1) 6 ppm parent ion mass tolerance, (2) partial tryptic digestion, (3) cysteine carbamidomethylation (+57.0215) and N-terminal/lysine 8-plex iTRAQ (+304.2053) addition as static modifications, and (4) oxidation (+15.9949 Da) on methionine, cysteine, tyrosine, and tryptophan, dioxidation (+31.9898 Da) on cysteine, and deamidation/deamination (+ 0.9840 Da) on asparagine, glutamine, and arginine residues as variable modifications. Identifications were filtered with MSGF probability scores of ≤1.0×10^−9^, ≤7×10^−11^ and ≤2×10^−12^ at spectral, peptide and protein levels, respectively, resulting in <1% false-discovery rate. iTRAQ reporter ion intensities were extracted with MASIC^41^, and the intensities of multiple MS/MS spectra from the same peptide were summed together to remove redundancy.

### Validation Phase - Targeted proteomics analysis

Up to 5 peptides were selected as surrogates for candidate biomarker proteins identified in the discovery phase based on their physical-chemical properties (between 8 and 20 amino acid residues, derived from trypsin digestion at both termini, and lack of post-translationally modified amino acid residues or residues that are problematic for chemical synthesis), and a Bayesian network-generated probabilistic score was used to select the peptides more likely to be successfully developed into targeted proteomics assays. To account for possible proteoforms, peptides from the same proteins that were not statistically significant were also included. Whole plasma of 6,426 individual samples were digested in batches of approximately 80 samples in 96-well plates^36^ and spiked with custom synthesized peptides (New England Peptides, now Vivitide) containing heavy isotopes in the C-terminal residues. Targeted proteomics analyses were performed using a Nano M-class UPLC (Waters) interfaced to a TSQ Altis triple quadrupole mass spectrometer (Thermo Fisher Scientific). Data were analyzed with the Skyline software and were manually inspected for proper alignment and background threshold.

### Statistical analysis

Statistical quality control of untargeted proteomics data involved removing peptides that were observed in only one sample per group and outlier identification using a Mahalanobis distance method^42,43^. Protein quantification from the peptide-level data was based on standard and scaled median quantification^43,44^ and statistics were performed on proteins and proteoforms (different forms of the same proteins resulting from gene isoforms, processing or post-translational modifications) based on their abundance profiles using an analysis of variance model, while accounting for sample pairing and batch in the model. The p-values were subsequently corrected with a Benjamini-Hochberg multiple comparison adjustment^45^ within each comparison to account for the multiple tests being performed. Machine learning was also performed to identify possible validation candidate proteins that did not meet the p-value threshold but were predictive of outcome in a multi-variate model. This was done using R and consisted of data imputation with Random Forest^46^, risk association via Probabilistic Conditional Logistic Regression integrated with least absolute shrinkage and selection operator (LASSO) for feature selection (clogitLasso)^47^.

### Machine learning analysis to identify early biomarker panels predictive of disease onset

Validation phase data was filtered to remove 3 peptides observed in less than 50% of samples for at least one of the three time points prior to seroconversion. Remaining missing values were imputed with Random Forest^46^ imputation. A pairing correction^48^ was applied to the data to account for the case-control study design. Logistic regression with a LASSO penalization function was fit to the data with case/control status as the explanatory variable. The machine learning model was fit separately to each time point’s data using four-fold cross-validation repeated for 100 bootstrap iterations.

### Function-enrichment analysis

Differentially abundant proteins were filtered for function-enrichment analysis using DAVID^49^, and only pathways containing KEGG annotation were used. The biological interpretations were only performed after the targeted proteomics data analysis were completed to avoid unconscious bias in sample and data analysis.

## Supporting information

List of TEDDY members

## Data Availability

All the collected data is available upon request at the TEDDY website.

https://teddy.epi.usf.edu/research/

## Authors’ contributions

E.S.N., C.A., M.A.G., A.A.S., L.M.B., P.D.P., W.J.Q, B.I.F, S.S., J.T., A.Z., A.L., W.H., B.A., R.D.S., B.J.M.W.R., M.J.R., T.O.M. and T.S.G conceived the study and participated in the study design. T.S.G. collected the samples. M.A.G., A.A.S., T.R.C., P.D.P., T.L.F., D.J.O. and R.J.M. prepared the samples and performed the LC-MS analyses. E.S.N., C.A., L.M.B., P.D.P., Y.G., B.A.S., D.J.O., W.J.Q, B.I.F, R.D.S., B.J.M.W.R., M.J.R., T.O.M. and T.S.G performed the data analysis. E.S.N., C.A., M.A.G., A.A.S., L.M.B., P.D.P., D.W.E., T.L.F., W.J.Q, R.D.S., B.J.M.W.R., M.J.R., T.O.M. and T.S.G developed the targeted mass spectrometry assays for validating the biomarker candidates. E.S.N., L.M.B, A.A.S., P.D.P., T.R.C., B.J.M.W.R., and T.O.M. wrote the manuscript. All authors read, revised, and approved the final manuscript for publication.

## Acknowledgments

The authors thank Mr. Nathan Johnson of PNNL for his help with Figures 1 and 6, and Drs. Jennifer Van Eyk (Cedars-Sinai Medical Center) and Michael McCoss (University of Washington) for insightful discussion. Part of the work was performed in the Environmental Molecular Sciences Laboratory, a U.S. DOE national scientific user facility at Pacific Northwest National Laboratory (PNNL) in Richland, WA. Battelle operates PNNL for the DOE under contract DE-AC05-76RLO01830.

## Data and Resource Availability

For data and resource availability, please reference the TEDDY data summary and access page: https://teddy.epi.usf.edu/research/.

## Supplementary Information

- **Supplementary Material**. Author information.
- **Supplementary information Table S1** – Statistical analysis of proteins quantified in the discovery phase.
- **Supplementary information Table S2 -** Biomarker candidates selected for the validation phase.
- **Supplementary information Table S3 -** Statistical analysis of peptides measured in the validation phase for individuals that developed type 1 diabetes.
- **Supplementary information Table S4** - Statistical analysis of peptides measured in the validation phase for individuals that had islet autoimmunity with normoglycemia.
- **Supplementary information Table S5** – Panels of peptides used for the multivariate panel prediction of islet autoimmunity with normoglycemia and type 1 diabetes development at the 6 months prior to seroconversion time point.

---

**Figure S1.**
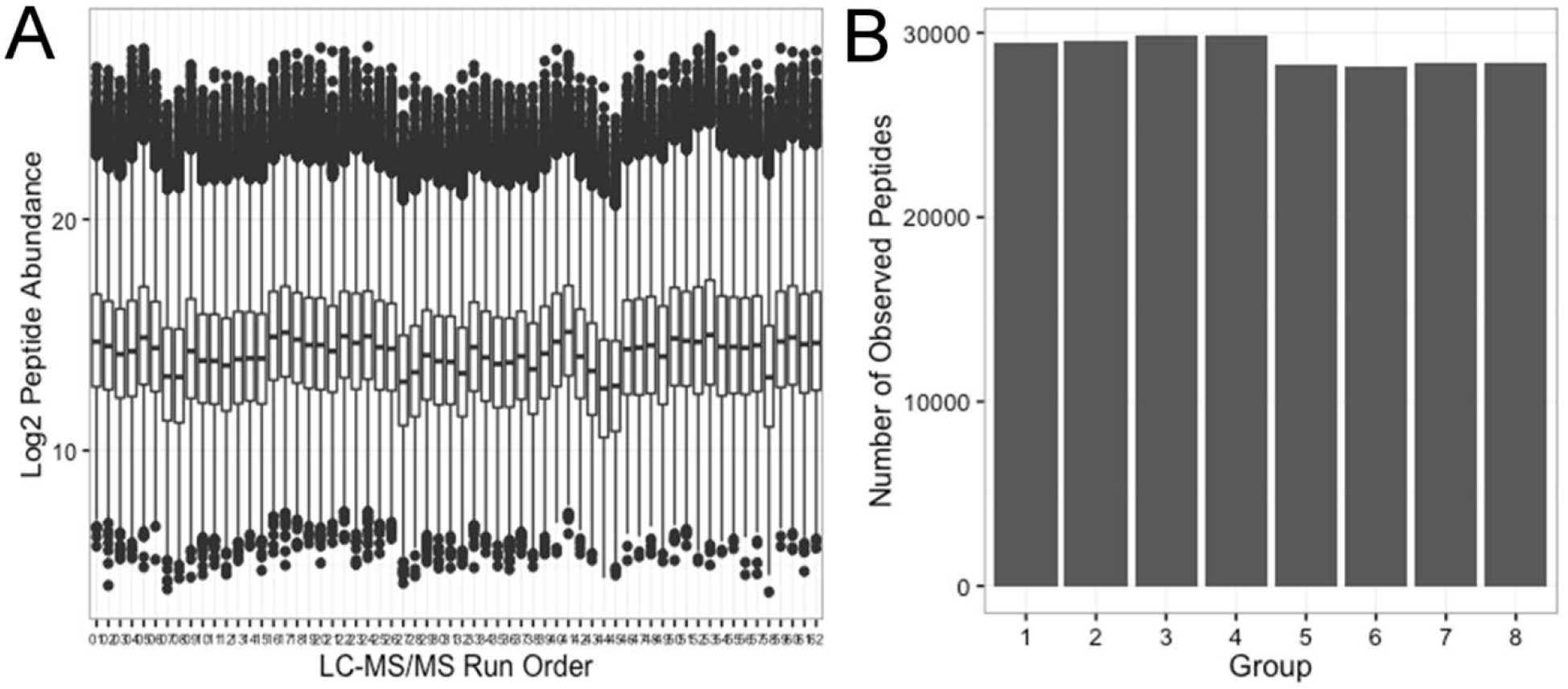
Data quality across the entire discovery phase analysis. (A) Overall peptide abundances of the reference sample channel in each of the sixty-two 8-plex iTRAQ sets. (B) Number of identified peptides in different sample groups.

**Figure S2.**
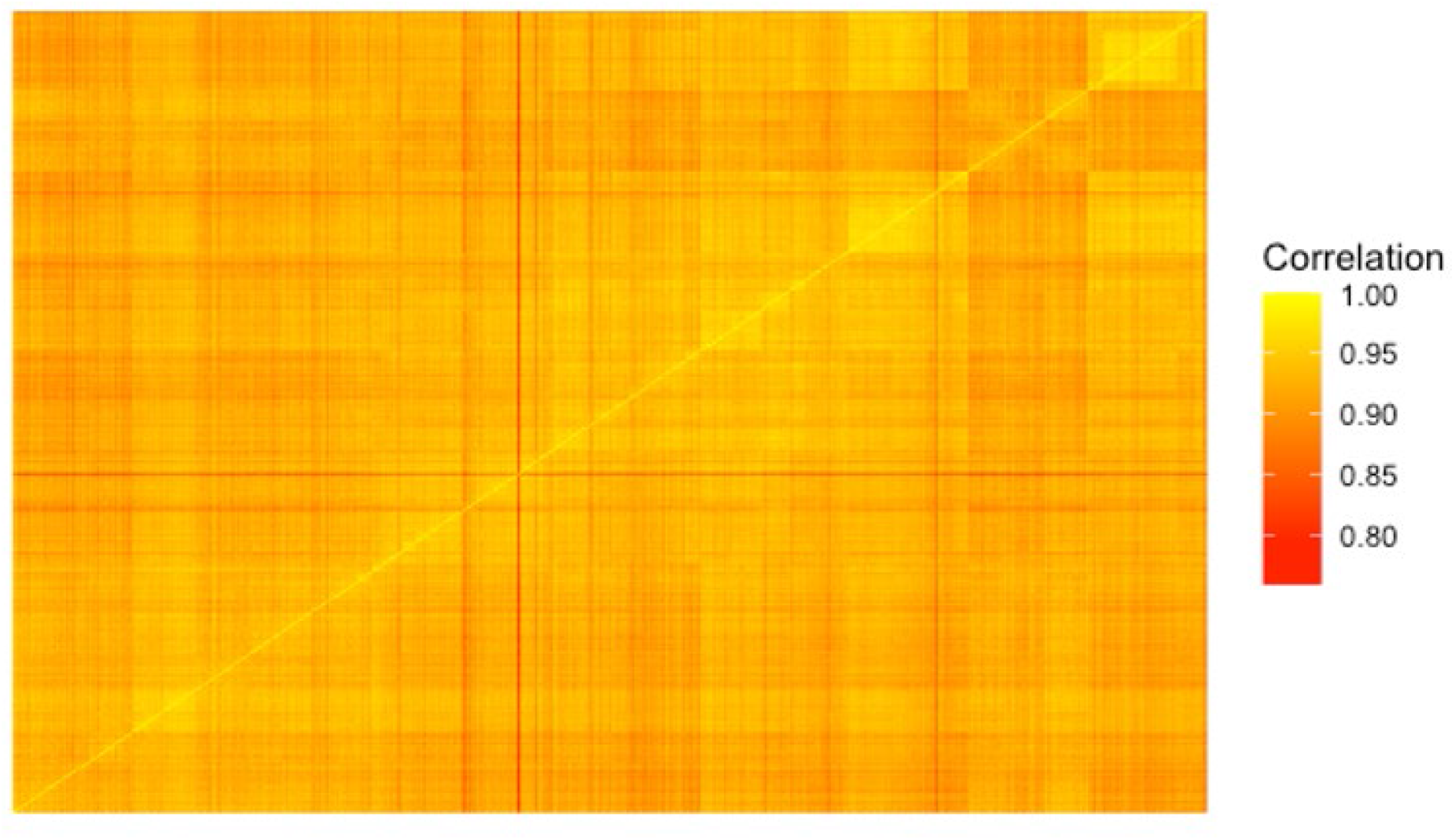
Data quality across the whole plasma proteomics analysis of the validation phase. The heatmap shows the correlations of the overall peptide abundances across the 6,426 targeted mass spectrometry analyses.

## References

1. Livingstone, S.J., et al. Estimated life expectancy in a Scottish cohort with type 1 diabetes, 2008-2010. JAMA 313, 37–44 (2015).

2. Atkinson, M.A., Eisenbarth, G.S. & Michels, A.W. Type 1 diabetes. Lancet 383, 69–82 (2014).

3. DiMeglio, L.A., Evans-Molina, C. & Oram, R.A. Type 1 diabetes. Lancet 391, 2449–2462 (2018).

4. Lee, H.S., et al. Biomarker discovery study design for type 1 diabetes in The Environmental Determinants of Diabetes in the Young (TEDDY) study. Diabetes Metab Res Rev 30, 424–434 (2014).

5. Keshishian, H., et al. Quantitative, multiplexed workflow for deep analysis of human blood plasma and biomarker discovery by mass spectrometry. Nat Protoc 12, 1683–1701 (2017).

6. Geyer, P.E., et al. Plasma Proteome Profiling to Assess Human Health and Disease. Cell Syst 2, 185–195 (2016).

7. Liu, Y., et al. Quantitative variability of 342 plasma proteins in a human twin population. Mol Syst Biol 11, 786 (2015).

8. Zhang, Q., et al. Serum proteomics reveals systemic dysregulation of innate immunity in type 1 diabetes. J Exp Med 210, 191–203 (2013).

9. Zhi, W., et al. Discovery and validation of serum protein changes in type 1 diabetes patients using high throughput two dimensional liquid chromatography-mass spectrometry and immunoassays. Mol Cell Proteomics 10, M111 012203 (2011).

10. von Toerne, C., et al. Peptide serum markers in islet autoantibody-positive children. Diabetologia 60, 287–295 (2017).

11. Moulder, R., et al. Serum proteomes distinguish children developing type 1 diabetes in a cohort with HLA-conferred susceptibility. Diabetes 64, 2265–2278 (2015).

12. MacLean, E., et al. A systematic review of biomarkers to detect active tuberculosis. Nat Microbiol 4, 748–758 (2019).

13. Nakayasu, E.S., et al. Tutorial: best practices and considerations for mass-spectrometry-based protein biomarker discovery and validation. Nat Protoc 16, 3737–3760 (2021).

14. Shi, T., et al. Antibody-free, targeted mass-spectrometric approach for quantification of proteins at low picogram per milliliter levels in human plasma/serum. Proc Natl Acad Sci U S A 109, 15395–15400 (2012).

15. Stanfill, B.A., et al. QC-ART: A tool for real-time quality control assessment of mass spectrometry-based proteomics data. Mol Cell Proteomics (2018).

16. Nakayasu, E.S., et al. Comprehensive Proteomics Analysis of Stressed Human Islets Identifies GDF15 as a Target for Type 1 Diabetes Intervention. Cell Metab 31, 363–374 e366 (2020).

17. Ramos-Rodriguez, M., et al. The impact of proinflammatory cytokines on the beta-cell regulatory landscape provides insights into the genetics of type 1 diabetes. Nat Genet 51, 1588–1595 (2019).

18. Gibbons, B.C., et al. Rapidly Assessing the Quality of Targeted Proteomics Experiments through Monitoring Stable-Isotope Labeled Standards. J Proteome Res 18, 694–699 (2019).

19. Marcelli-Barge, A., et al. Marked shortage of C4B DNA polymorphism among insulin-dependent diabetic patients. Res Immunol 141, 117–128 (1990).

20. Torn, C., et al. Complement gene variants in relation to autoantibodies to beta cell specific antigens and type 1 diabetes in the TEDDY Study. Sci Rep 6, 27887 (2016).

21. Rowe, P., et al. Increased complement activation in human type 1 diabetes pancreata. Diabetes Care 36, 3815–3817 (2013).

22. Targher, G., Chonchol, M., Zoppini, G. & Franchini, M. Hemostatic disorders in type 1 diabetes mellitus. Semin Thromb Hemost 37, 58–65 (2011).

23. Nejentsev, S., et al. Localization of type 1 diabetes susceptibility to the MHC class I genes HLA-B and HLA-A. Nature 450, 887–892 (2007).

24. van Lummel, M., et al. Discovery of a Selective Islet Peptidome Presented by the Highest-Risk HLA-DQ8trans Molecule. Diabetes 65, 732–741 (2016).

25. Eizirik, D.L., Colli, M.L. & Ortis, F. The role of inflammation in insulitis and beta-cell loss in type 1 diabetes. Nat Rev Endocrinol 5, 219–226 (2009).

26. Webb-Robertson, B.M., et al. Prediction of the development of islet autoantibodies through integration of environmental, genetic, and metabolic markers. J Diabetes 13, 143–153 (2021).

27. Webb-Robertson, B.M., et al. Integration of Infant Metabolite, Genetic and Islet Autoimmunity Signatures to Predict Type 1 Diabetes by 6 Years of Age. J Clin Endocrinol Metab (2022).

28. Sims, E.K., et al. Elevations in the Fasting Serum Proinsulin-to-C-Peptide Ratio Precede the Onset of Type 1 Diabetes. Diabetes Care 39, 1519–1526 (2016).

29. Lamichhane, S., et al. Dynamics of Plasma Lipidome in Progression to Islet Autoimmunity and Type 1 Diabetes - Type 1 Diabetes Prediction and Prevention Study (DIPP). Sci Rep 8, 10635 (2018).

30. Valladolid-Acebes, I., Berggren, P.O. & Juntti-Berggren, L. Apolipoprotein CIII Is an Important Piece in the Type-1 Diabetes Jigsaw Puzzle. Int J Mol Sci 22(2021).

31. Morillas, P., et al. Circulating biomarkers of collagen metabolism in arterial hypertension: relevance of target organ damage. J Hypertens 31, 1611–1617 (2013).

32. Medina, C.O., Nagy, N. & Bollyky, P.L. Extracellular matrix and the maintenance and loss of peripheral immune tolerance in autoimmune insulitis. Curr Opin Immunol 55, 22–30 (2018).

33. Lu, G., et al. Dextran Sulfate Protects Pancreatic beta-Cells, Reduces Autoimmunity, and Ameliorates Type 1 Diabetes. Diabetes 69, 1692–1707 (2020).

34. Mathieu, C., Lahesmaa, R., Bonifacio, E., Achenbach, P. & Tree, T. Immunological biomarkers for the development and progression of type 1 diabetes. Diabetologia 61, 2252–2258 (2018).

35. Ahmed, S., et al. Standardizing T-Cell Biomarkers in Type 1 Diabetes: Challenges and Recent Advances. Diabetes 68, 1366–1379 (2019).

36. Piehowski, P.D., et al. Sources of technical variability in quantitative LC-MS proteomics: human brain tissue sample analysis. J Proteome Res 12, 2128–2137 (2013).

37. Gritsenko, M.A., Xu, Z., Liu, T. & Smith, R.D. Large-Scale and Deep Quantitative Proteome Profiling Using Isobaric Labeling Coupled with Two-Dimensional LC-MS/MS. Methods Mol Biol 1410, 237–247 (2016).

38. Mayampurath, A.M., et al. DeconMSn: a software tool for accurate parent ion monoisotopic mass determination for tandem mass spectra. Bioinformatics 24, 1021–1023 (2008).

39. Petyuk, V.A., et al. DtaRefinery, a software tool for elimination of systematic errors from parent ion mass measurements in tandem mass spectra data sets. Mol Cell Proteomics 9, 486–496 (2010).

40. Kim, S. & Pevzner, P.A. MS-GF+ makes progress towards a universal database search tool for proteomics. Nat Commun 5, 5277 (2014).

41. Monroe, M.E., Shaw, J.L., Daly, D.S., Adkins, J.N. & Smith, R.D. MASIC: a software program for fast quantitation and flexible visualization of chromatographic profiles from detected LC-MS(/MS) features. Comput Biol Chem 32, 215–217 (2008).

42. Matzke, M.M., et al. Improved quality control processing of peptide-centric LC-MS proteomics data. Bioinformatics 27, 2866–2872 (2011).

43. Webb-Robertson, B.J., et al. Bayesian proteoform modeling improves protein quantification of global proteomic measurements. Mol Cell Proteomics 13, 3639–3646 (2014).

44. Matzke, M.M., et al. A comparative analysis of computational approaches to relative protein quantification using peptide peak intensities in label-free LC-MS proteomics experiments. Proteomics 13, 493–503 (2013).

45. Benjamini, Y. & Hochberg, Y. Controlling the False Discovery Rate: A Practical and Powerful Approach to Multiple Testing Journal of the Royal Statistical Society. Series B (Methodological) 57, 289–300 (1995).

46. Liaw, A. & Wiener, M. Classification and Regression by Random Forest. R News 2, 18–22 (2002).

47. Avalos, M., Pouyes, H., Grandvalet, Y., Orriols, L. & Lagarde, E. Sparse conditional logistic regression for analyzing large-scale matched data from epidemiological studies: a simple algorithm. BMC Bioinformatics 16 Suppl 6, S1 (2015).

48. Stanfill, B., et al. Extending Classification Algorithms to Case-Control Studies. Biomed Eng Comput Biol 10, 1179597219858954 (2019).

49. Huang da, W., Sherman, B.T. & Lempicki, R.A. Bioinformatics enrichment tools: paths toward the comprehensive functional analysis of large gene lists. Nucleic Acids Res 37, 1–13 (2009).

