## Supplementary material for "Plasma protein biomarkers predict both the development of persistent autoantibodies and type 1 diabetes 6 months prior to the onset of autoimmunity: the TEDDY Study": List of TEDDY members

**The TEDDY Study Group**

**Colorado Clinical Center:** Marian Rewers, M.D., Ph.D., PI^1,4,6,9,10^, Kimberly Bautista^11^, Judith Baxter^8,911^, Daniel Felipe-Morales, Brigitte I. Frohnert, M.D., Ph.D.^2,13^, Marisa Stahl, M.D.^12^, Isabel Flores Garcia, Patricia Gesualdo^2,6,11,13^, Sierra Hays, Michelle Hoffman^11,12,13^, Rachel Karban^11^, Edwin Liu, M.D.^12^, Leila Loaiza Jill Norris, Ph.D.^2,3,11^, Holly O’Donnell, Ph.D.^8^, Loana Thorndahl, Andrea Steck, M.D.^3,13^, Kathleen Waugh^6,7,11^. University of Colorado, Anschutz Medical Campus, Barbara Davis Center for Childhood Diabetes, Aurora, CO, USA.

**Finland Clinical Center:** Jorma Toppari, M.D., Ph.D., PI^¥^1,4,10,13^, Olli G. Simell, M.D., Ph.D., Annika Adamsson, Ph.D.^^11^, Suvi Ahonen*^±§^, Mari Åkerlund*^±§^, Sirpa Anttila^µ¤^, Leena Hakola*^±^, Anne Hekkala, M.D.^µ¤^, Tiia Honkanen^µ¤^, Heikki Hyöty, M.D., Ph.D.*^±6^, Jorma Ilonen, M.D., Ph.D.^¥3^, Sanna Jokipuu^^^, Taru Karjalainen^µ¤^, Leena Karlsson^^^, Jukka Kero M.D., Ph.D.^¥^3, 13^, Jaakko J. Koskenniemi M.D., Ph.D.^¥^^, Miia Kähönen^µ¤11,13^, Mikael Knip, M.D., Ph.D.*^±^, Minna-Liisa Koivikko^µ¤^, Katja Kokkonen*^±^, Merja Koskinen*^±^, Mirva Koreasalo*^±§2^, Kalle Kurppa, M.D., Ph.D.*^±12^, Salla Kuusela, M.D. ^µ¤^, Jarita Kytölä^*±^, Jutta Laiho, Ph.D.*^6^, Tiina Latva-aho^µ¤^, Siiri Leisku^*±^, Laura Leppänen^^^, Katri Lindfors, Ph.D.*^12^, Maria Lönnrot, M.D., Ph.D.*^±6^, Elina Mäntymäki^^^, Markus Mattila*^±^, Maija Miettinen^§2^, Teija Mykkänen^µ¤^, Tiina Niininen^±^*^11^, Sari Niinistö^§2^, Noora Nurminen^*±^, Sami Oikarinen, Ph.D.*^±6^, Hanna-Leena Oinas*^±^, Paula Ollikainen^µ¤^, Zhian Othmani^¥^, Sirpa Pohjola ^µ¤^, Solja Raja-Hanhela^µ¤^, Jenna Rautanen^±§^, Anne Riikonen*^±§2^, Minna Romo^^^, Juulia Rönkä^µ¤^, Nelli Rönkä^µ¤^, Satu Simell, M.D., Ph.D.^¥12^, Päivi Tossavainen, M.D.^µ¤^, Mari Vähä-Mäkilä^¥^, Eeva Varjonen^^11^, Riitta Veijola, M.D., Ph.D.^µ¤13^, Irene Viinikangas^µ¤^, Silja Vilmi^µ¤^, Suvi M. Virtanen, M.D., Ph.D.*^±§2^. ^¥^University of Turku, Turku, Finland, *Tampere University, Tampere, Finland, ^µ^University of Oulu, Oulu, Finland, ^^^Turku University Hospital, Hospital District of Southwest Finland, Turku, Finland, ^±^Tampere University Hospital, Tampere, Finland, ^¤^Oulu University Hospital, Oulu, Finland, §Finnish Institute for Health and Welfare, Helsinki, Finland.

**Georgia/Florida Clinical Center:** Richard McIndoe, Ph.D., PI^^4,10^, Desmond Schatz, M.D.*^4,7,8^, Diane Hopkins^^11^, Michael Haller, M.D.*^13^, Risa Bernard^^11^, Melissa Gardiner^^11^, Ashok Sharma, Ph.D.^^^, Laura Jacobsen, M.D.*^13^, Jennifer Hosford^^^, Kennedy Petty^^^, Leah Myers^^^, Chelsea Salmon*. ^^^Center for Biotechnology and Genomic Medicine, Augusta University, Augusta, GA, USA. *University of Florida, Pediatric Endocrinology, Gainesville, FL, USA.

**Germany Clinical Center:** Anette G. Ziegler, M.D., PI^1,3,4,10^, Ezio Bonifacio Ph.D.*, Cigdem Gezginci, Willi Grätz, Anja Heublein, Eva Hohoff^¥2^, Sandra Hummel, Ph.D.^2^, Annette Knopff^7^, Melanie Köger, Sibylle Koletzko, M.D.^¶12^, Claudia Ramminger^11^, Roswith Roth, Ph.D.^8^, Jennifer Schmidt, Marlon Scholz, Joanna Stock^8,11,13^, Katharina Warncke, M.D.^13^, Lorena Wendel, Christiane Winkler, Ph.D.^2,11^. Forschergruppe Diabetes e.V. and Institute of Diabetes Research, Helmholtz Zentrum München, Forschergruppe Diabetes, and Klinikum rechts der Isar, Technische Universität München, Neuherberg, Germany. *Center for Regenerative Therapies, TU Dresden, Dresden, Germany, ^¶^Dr. von Hauner Children’s Hospital, Department of Gastroenterology, Ludwig Maximillians University Munich, Munich, Germany, ^¥^University of Bonn, Department of Nutritional Epidemiology, Bonn, Germany.

**Sweden Clinical Center:** Åke Lernmark, Ph.D., PI^1,3,4,5,6,8,9,10^, Daniel Agardh, M.D., Ph.D.^6,12^, Carin Andrén Aronsson, Ph.D.^2,11,12^, Rasmus Bennet, Corrado Cilio, Ph.D., M.D.^6^, Susanne Dahlberg, Ulla Fält, Malin Goldman Tsubarah, Emelie Ericson-Hallström, Lina Fransson, Emina Halilovic, Gunilla Holmén, Susanne Hyberg, Berglind Jonsdottir, M.D., Ph.D.^11^, Naghmeh Karimi, Helena Elding Larsson, M.D., Ph.D.^6,13^, Marielle Lindström, Markus Lundgren, M.D., Ph.D.^13^, Marlena Maziarz, Ph.D., Jessica Melin^11^, Caroline Nilsson, Kobra Rahmati, Anita Ramelius, Falastin Salami, Ph.D., Anette Sjöberg, Evelyn Tekum Amboh Carina Törn, Ph.D.^3^, Ulrika Ulvenhag, Terese Wiktorsson, Åsa Wimar^13^. Lund University, Lund, Sweden.

**Washington Clinical Center:** William A. Hagopian, M.D., Ph.D., PI^1,3,4,6,7,10,12,13^, Michael Killian^6,7,11,12^, Claire Cowen Crouch^11,13^, Jennifer Skidmore^2^, Trevor Bender, Megan Llewellyn, Cody McCall, Arlene Meyer, Jocelyn Meyer, Denise Mulenga^11^, Nole Powell, Jared Radtke, Shreya Roy, Preston Tucker. Pacific Northwest Research Institute, Seattle, WA, USA.

**Pennsylvania Satellite Center:** Dorothy Becker, M.D., Margaret Franciscus, MaryEllen Dalmagro-Elias Smith^2^, Ashi Daftary, M.D., Mary Beth Klein, Chrystal Yates. Children’s Hospital of Pittsburgh of UPMC, Pittsburgh, PA, USA.

**Data Coordinating Center:** Jeffrey P. Krischer, Ph.D., PI^1,4,5,9,10^, Rajesh Adusumali, Sarah Austin-Gonzalez, Maryouri Avendano, Sandra Baethke, Brant Burkhardt, Ph.D.^6^, Martha Butterworth^2^, Nicholas Cadigan, Joanna Clasen, Kevin Counts, Laura Gandolfo, Jennifer Garmeson, Veena Gowda, Christina Karges, Shu Liu, Xiang Liu, Ph.D.^2,3,8,13^, Kristian Lynch, Ph.D. ^6,8^, Jamie Malloy, Lazarus Mramba, Ph.D.^2^, Cristina McCarthy^11^, Jose Moreno, Hemang M. Parikh, Ph.D.^3,8^, Cassandra Remedios, Chris Shaffer, Susan Smith^11^, Noah Sulman, Ph.D., Roy Tamura, Ph.D.^1,2,11,12,13^, Dena Tewey, Henri Thuma, Michael Toth, Ulla Uusitalo, Ph.D.^2^, Kendra Vehik, Ph.D.^4,5,6,8,13^, Ponni Vijayakandipan, Melissa Wroble, Jimin Yang, Ph.D., R.D.^2^, Kenneth Young, Ph.D. *Past staff: Michael Abbondondolo, Lori Ballard, Rasheedah Brown, David Cuthbertson, Stephen Dankyi, Christopher Eberhard, Steven Fiske, David Hadley, Ph.D., Kathleen Heyman, Belinda Hsiao, Francisco Perez Laras, Hye-Seung Lee, Ph.D., Qian Li, Ph.D., Colleen Maguire, Wendy McLeod, Aubrie Merrell, Steven Meulemans, Ryan Quigley, Laura Smith, Ph.D.* University of South Florida, Tampa, FL, USA.

**Project scientist:** Beena Akolkar, Ph.D.^1,3,4,5,6,7,9,10^. National Institutes of Diabetes and Digestive and Kidney Diseases, Bethesda, MD, USA.

**Proteomics Laboratory:** Richard D. Smith, Ph.D., Thomas O. Metz, Ph.D., Bobbie-Jo Webb-Robertson, Ph.D., Paul D. Piehowski, Ph.D., Ernesto S. Nakayasu, Ph.D., Lisa Bramer, Ph.D., and Wei-Jun Qian, Ph.D. Pacific Northwest National Laboratory, Richland, WA, USA.

**Repository:** Chris Deigan. NIDDK Biosample Repository at Fisher BioServices, Rockville, MD, USA. (Previously Ricky Schrock, Polina Malone, Sandra Ke, Niveen Mulholland, Ph.D.)

**Other contributors:** Thomas Briese, Ph.D.^6^, Columbia University. Todd Brusko, Ph.D.^5^, University of Florida, Gainesville, FL, USA. Teresa Buckner, Ph.D.^2^, University of Northern Colorado, Greeley, CO. Suzanne Bennett Johnson, Ph.D.^8,11^, Florida State University, Tallahassee, FL, USA. Eoin McKinney, Ph.D.^5^, University of Cambridge, Cambridge, UK. Tomi Pastinen, M.D., Ph.D.^5^, The Children’s Mercy Hospital, Kansas City, MO, USA. Steffen Ullitz Thorsen, M.D., Ph.D.^2^, Department of Clinical Immunology, University of Copenhagen, Copenhagen, Denmark, and Department of Pediatrics and Adolescents, Copenhagen University Hospital, Herlev, Denmark. Eric Triplett, Ph.D.^6^, University of Florida, Gainesville, FL, USA.

***Committees:***

^1^Ancillary Studies, ^2^Diet, ^3^Genetics, ^4^Human Subjects/Publicity/Publications, ^5^Immune Markers, ^6^Infectious Agents, ^7^Laboratory Implementation, ^8^Psychosocial, ^9^Quality Assurance, ^10^Steering, ^11^Study Coordinators, ^12^Celiac Disease, ^13^Clinical Implementation.
